# ADataViewer: Exploring Semantically Harmonized Alzheimer’s Disease Cohort Datasets

**DOI:** 10.1101/2021.09.01.21262607

**Authors:** Yasamin Salimi, Daniel Domingo-Fernándéz, Carlos Bobis-Álvarez, Martin Hofmann-Apitius, for the Alzheimer’s Disease Neuroimaging Initiative, the Japanese Alzheimer’s Disease Neuroimaging Initiative, for the Aging Brain: Vasculature, Ischemia, and Behavior Study, the Alzheimer’s Disease Repository Without Borders Investigators, for the European Prevention of Alzheimer’s Disease (EPAD) Consortium, Colin Birkenbihl

## Abstract

**INTRODUCTION:** Currently, AD cohort datasets are difficult to find, lack across-cohort interoperability, and the content of the shared datasets often only becomes clear to third-party researchers once data access has been granted.

**METHODS:** We accessed and systematically investigated the content of 20 major AD cohort datasets on data-level. A medical professional and a data specialist manually curated and semantically harmonized the acquired datasets. We developed a platform that facilitates data exploration.

**RESULTS:** We present ADataViewer, an interactive platform that facilitates the exploration of 20 cohort datasets with respect to longitudinal follow-up, demographics, ethnoracial diversity, measured modalities, and statistical properties of individual variables. Additionally, we publish a variable mapping catalog harmonizing 1,196 variables across the 20 cohorts. The platform is available under https://adata.scai.fraunhofer.de/.

**DISCUSSION:** ADataViewer supports robust data-driven research by transparently displaying cohort dataset content and suggesting datasets suited for discovery and validation studies based on selected variables of interest.

## 1. Background

Alzheimer’s disease (AD) and dementia research has progressed considerably thanks to the increased availability of patient-level cohort datasets [1]. Cohort data have, among others, laid the foundation to discover novel biomarkers [2], investigate disease progression [3], and identify disease subtypes [4]. To ensure the robustness and reproducibility of results achieved in such data-driven analyses, it is crucial that they are externally validated in independent cohort datasets [5]. Working across multiple cohort datasets is, however, impeded by several profound challenges. The first challenge manifests in the access to further validation cohort datasets, as third-party researchers have to go through time-intensive application processes that often span several weeks before they can actually start getting familiar with the acquired data. Secondly, once access is granted, the validation datasets have to be comparable to the original discovery dataset with respect to their assessed variables [6]. This means that 1) a largely overlapping set of variables should have been measured in both cohorts, and 2) these variables need to be harmonized across the independent cohort datasets, which is rarely the case by default. Identifying and semantically harmonizing equivalent variables in distinct datasets is an arduous task given that datasets typically employ their own variable naming system [7]. While theoretical guidelines for AD data harmonization have been previously proposed [8], as of now and to the best of our knowledge, no comprehensive mapping catalog is available to the AD research community that would help to unify the variable names across existing cohorts.

Across-cohort interoperability, however, goes beyond the semantic layer as statistical distributions of equivalent variables might differ among cohorts [9]. Our recent study revealed that such systematic statistical differences can bias results of data-driven analyses based on cohort data [10]. However, in practice, researchers only see the factual content of a shared dataset after data download occurred and data investigation started. At this stage, the realization of incompatible discovery and validation datasets can render the process of data access and exploration a waste of time.

Several funding bodies, for example the Innovative Medicine Initiative (IMI) or the Alzheimer’s Disease Data Initiative (ADDI), have launched large projects to address these problems, such as the European Medical Information Framework (EMIF) [11], ROADMAP [12] or the ADDI Workbench, and new calls were issued in this direction. In fact, both EMIF and ROADMAP have built information sources on cohort datasets that were assembled from the respective cohorts’ self-reported metadata [13,14]. However, in a recent study, we observed that the information gained through such metadata-driven cohort assessments differs from the content that is factually shared with researchers after successful access applications [15].

In this work, we present ADataViewer, an interactive tool that enables the scientific community to explore 20 AD cohort datasets, both from a semantic and statistical perspective. To establish semantic interoperability across these datasets, we created a variable mapping catalog which harmonizes 1,196 unique variables encountered in the datasets, spanning nine data modalities. Leveraging these semantically harmonized versions of the datasets, we developed tools and interfaces that facilitate the exploration of the cohort datasets with respect to longitudinal follow-up, demographics, ethnoracial diversity, measured modalities, and individual variables. Finally, we present ADataViewers’ ‘StudyPicker’, a tool that assists researchers in identifying cohort datasets suited for their envisioned analysis.

## 2. Methods

### 2.1 AD cohort datasets

To enable a comprehensive exploration of the available data in the AD domain, it was vital to identify, access and curate as many cohort-level datasets as possible. Therefore, we systematically scanned data repositories and scientific publications, leading to the identification of 24 cohorts that claimed to follow the open science paradigm and share their data with third-party researchers. After applying for access to the corresponding data owners, we acquired 20 of those datasets over the course of three years. These datasets originated from a heterogeneous pool of studies that followed a variety of different goals ranging from purely observational cohort studies over memory clinic data collections to dedicated clinical trials. Concordantly, the employed participant recruitment procedures, inclusion and exclusion criteria, and measured data modalities varied among them. More information about the collected datasets, their content and original study aim are given in **Table 1**; for further study-specific details we refer to the original publications.

**Table 1.** The investigated AD cohorts available for exploration through the ADataViewer.

| Cohort | Consortium | Patients at baseline | Modalities | Longitudinal (yes/no) | Study type |
| --- | --- | --- | --- | --- | --- |
| A4 [16] | Anti-Amyloid Treatment in Asymptomatic Alzheimer's Disease | 6,945 | 7 | No* | Clinical Trial |
| ABVIB [17] | Aging Brain: Vasculature, Ischemia, and Behavior | 280 | 2 | Yes | Observational Study |
| ADNI [18] | The Alzheimer's Disease Neuroimaging Initiative | 2,249 | 12 | Yes | Observational Study |
| AIBL [19] | The Australian Imaging, Biomarker & Lifestyle Flagship Study of Ageing | 1,378 | 9 | Yes | Observational Study |
| ANMerge [20] | AddNeuroMed | 1,703 | 10 | Yes | Observational Study |
| ARWIBO [21] | Alzheimer's Disease Repository Without Borders | 2,617 | 10 | Yes | Observational Study |
| DOD ADNI [22] | Effects of TBI & PTSD on Alzheimer's Disease in Vietnam Vets | 458 | 11 | Yes | Observational Study |
| EDSD [23] | The European DTI Study on Dementia | 474 | 7 | No | Observational Study |
| EMIF-1000 [24] | European Medical Information Framework | 1,199 | 10 | No | Meta-cohort |
| EPAD V.IMI [25] | European Prevention of Alzheimer's Dementia | 2,096 | 9 | Yes | Observational Study |
| I-ADNI [26] | The Italian Alzheimer's Disease Neuroimaging Initiative | 262 | 5 | No | Observational Study |
| JADNI [27] | Japanese Alzheimer's Disease Neuroimaging Initiative | 567 | 9 | Yes | Observational Study |
| NACC [28] | The National Alzheimer's Coordinating Center | 40,948 | 11 | Yes | Memory Clinic Database |
| OASIS [29] | Open Access Series of Imaging Studies | 564 | 3 | Yes | Observational Study |
| PREVENT-AD [30] | Pre-symptomatic Evaluation of Experimental or Novel Treatments for Alzheimer's Disease | 348 | 8 | Yes | Clinical Trial |
| PharmaCog [31] | Prediction of Cognitive Properties of New Drug Candidates for Neurodegenerative Diseases in Early Clinical Development | 147 | 6 | Yes | Observational Study |
| ROSMAP [32] | The Religious Orders Study and Memory and Aging Project | 3,626 | 7 | Yes | Observational Study |
| VASCULAR [33] | The Vascular Contributors to Prodromal Alzheimer's disease | 250 | 8 | No | Non-interventional Cohort Study |
| VITA [34] | Vienna Transdanube Aging | 606 | 5 | Yes | Observational Study |
| WMH-AD [35] | White Matter Hyperintensities in Alzheimer's Disease | 90 | 5 | No | Observational Study |

**Note:** A complete overview about the collected data modalities can be found under https://adata.scai.fraunhofer.de/modality. \*, Follow-up assessments were planned for A4 but no according data was released at the time of this publication.

### 2.2 Systematic assessment of cohort dataset content

We based all our investigations on the data that were factually shared by the respective data owners, instead of relying solely on study protocols and reported metadata. To transparently mirror the state of the dataset to which researchers will gain access after successful application, we refrained from any extensive data processing (e.g., transforming numerical ranges and value representations). As such, any inconsistencies in the datasets (e.g., extreme outliers) will be accordingly displayed in ADataViewers’ tools and visualizations. Consequently, this allows researchers to comprehensively evaluate the data that will actually be available for analysis.

### 2.3 Harmonizing variables across cohorts

Semantic harmonization of the datasets was achieved through meticulous manual curation. Two curators systematically investigated variable names, metadata describing the variable content, and the values stored in the respective data tables to gain robust mappings between equivalent variables. We opted for a multidisciplinary curation team to combine the complementary strengths of a curator from a medical background with those of a second curator leveraging a data-driven perspective. For more detailed curation guidelines, we refer to the **Supplementary Material**. Whenever possible, variables found in the investigated AD datasets were additionally mapped to ontologies that provided respective semantic context. Further details on the used ontologies and the process of mapping variable names to ontologies are described in the **Supplementary Material**.

### 2.4 Data access and data privacy

ADataViewer does not store or enable the download of any cohort data itself. All displayed plots and provided exploration tools are fully anonymized and no participant identifying information is disclosed nor stored in the underlying database, not even the original study internal patient identifiers. Shown statistical plots are solely based on summary statistics or univariate analyses that can not be linked to other variables or personal information. In order to facilitate access to the datasets, we provide links that lead researchers to the original data portals through which the respective cohorts are distributed.

## 3. Results

ADataViewer is an interactive platform that enables the detailed exploration of, at the time of publication, 20 major cohort datasets from the AD domain. Its goal is to provide an overview across their content from a predominantly data-driven perspective. Each section of ADataViewer focuses on distinct aspects of the investigated datasets. The ‘Modality’ section provides an overview on the data modalities collected in each cohort (e.g. magnetic resonance imaging (MRI), autopsy, and genotype data). The ‘Ethnicity’ page displays the ethnoracial diversity in each cohort study as well as aggregated plots over specific geographic regions. In the ‘Longitudinal’ section, the frequency and abundance of follow-up assessments is presented both per cohort and variable. The ‘Biomarkers’ section allows the visualization of variable distributions and their comparison across cohorts. The semantic mappings between cohort name spaces are covered in the ‘Mappings’ section. Finally, the ‘StudyPicker’ leverages on all of these sections to guide researchers to the cohort datasets which provide the best basis for their planned analyses.

### 3.1 Semantic harmonization of the accessed cohort datasets

To build ADataViewer, we mapped 1,196 unique terms across the investigated datasets corresponding to variables from nine different data modalities. **Table 2** shows the total number of mapped terms per modality and cohort. For the majority of modalities, we mapped approximately between 10 and 30 variables **(Fig. 1)**, with the exception being the MRI modality which contained over 1,000 variables, as it comprised a vast selection of brain region specific measures derived from the raw images (e.g. volumes or thickness). Furthermore, to connect the variables of the cohort datasets to clearly defined semantic concepts, we additionally mapped them to standardized ontologies. In total, 241 concepts from seven distinct referential ontologies were used in this process **(more details in the Supplements)**. All mappings can be explored through interactive visualizations and tables under https://adata.scai.fraunhofer.de/mappings. The genotype and omics modalities of datasets were not mapped as they are already precisely defined by genetic database identifiers (e.g. rsID’s or UniProt identifiers) and their corresponding reference genome.

**Table 2:** Summary statistics describing the semantic harmonization of cohort datasets.

| Dataset | Demographics | Clinical | MRI | PET | CSF | Plasma | Comorbidities | Family | Lifestyle |
| --- | --- | --- | --- | --- | --- | --- | --- | --- | --- |
| A4 | 13 | 5 | 44 | 1 | 0 | 0 | 2 | 6 | 4 |
| ABVIB | 12 | 0 | 0 | 0 | 0 | 0 | 0 | 0 | 0 |
| ADNI | 17 | 23 | 247 | 3 | 10 | 11 | 14 | 8 | 5 |
| AIBL | 15 | 16 | 3 | 2 | 3 | 0 | 12 | 2 | 5 |
| ANMerge | 14 | 11 | 136 | 0 | 0 | 0 | 1 | 3 | 1 |
| ARWIBO | 21 | 14 | 1,026 | 21 | 3 | 6 | 13 | 3 | 2 |
| DOD ADNI | 21 | 20 | 249 | 1 | 3 | 0 | 18 | 6 | 6 |
| EDSD | 12 | 8 | 1,026 | 8 | 3 | 2 | 4 | 2 | 0 |
| EMIF-1000 | 8 | 4 | 3 | 1 | 6 | 0 | 3 | 0 | 4 |
| EPAD V.IMI | 14 | 11 | 80 | 0 | 3 | 0 | 17 | 5 | 4 |
| I-ADNI | 15 | 10 | 1,026 | 8 | 3 | 1 | 1 | 2 | 0 |
| JADNI | 15 | 21 | 871 | 2 | 3 | 0 | 14 | 6 | 4 |
| NACC | 20 | 17 | 123 | 2 | 3 | 0 | 14 | 3 | 6 |
| OASIS | 16 | 3 | 1,026 | 8 | 3 | 2 | 0 | 2 | 0 |
| PREVENT-AD | 15 | 4 | 0 | 0 | 7 | 0 | 5 | 5 | 0 |
| PharmaCog | 13 | 16 | 1,026 | 8 | 3 | 2 | 0 | 2 | 0 |
| ROSMAP | 12 | 9 | 0 | 0 | 0 | 0 | 8 | 0 | 1 |
| VASCULAR | 9 | 8 | 31 | 0 | 0 | 0 | 3 | 0 | 2 |
| VITA | 12 | 3 | 1,026 | 8 | 3 | 2 | 0 | 2 | 0 |
| WMH-AD | 12 | 4 | 1,025 | 8 | 3 | 2 | 0 | 2 | 0 |
| Total unique terms | 23 | 34 | 1,050 | 24 | 14 | 15 | 20 | 9 | 7 |

**Figure 1:**
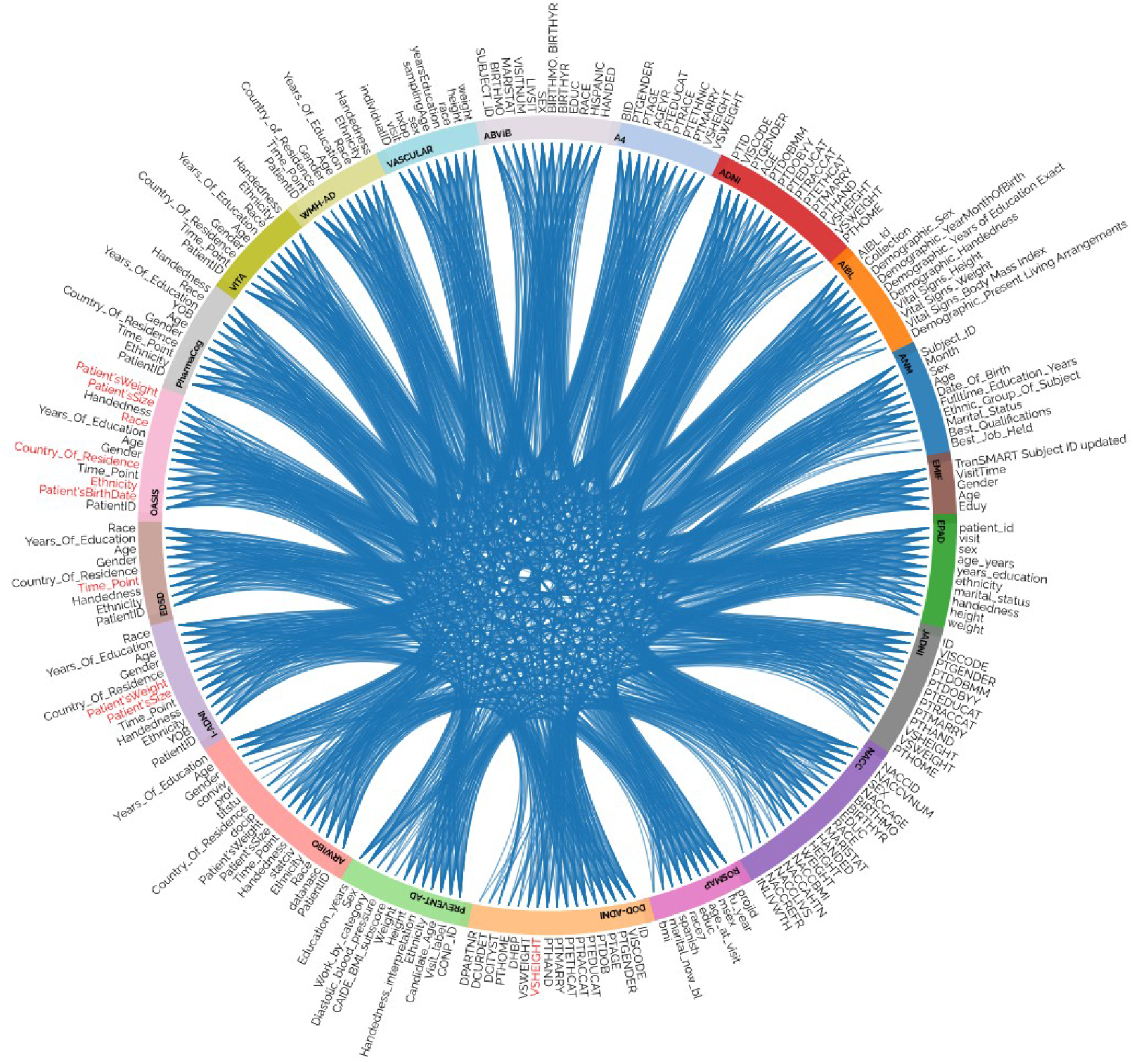
Circleplot presenting the mappings of demographic variables across the 20 cohorts. Red labels indicate variables mentioned in the metadata which consisted purely of missing data in the shared dataset. The corresponding plot for each modality as well as the underlying mapping tables for data harmonization are available at https://adata.scai.fraunhofer.de/mappings.

### 3.2 The StudyPicker: variable-based selection of cohort datasets

The StudyPicker is a tool that supports researchers in finding datasets based on the requirements of their envisioned analysis (https://adata.scai.fraunhofer.de/study_picker). It takes a collection of variable names as input and ranks the cohorts in ADataViewer based on the availability of these specified variables **(Fig. 4A)**. The generated ranking shows the availability of the variables, the number of participants per cohort for whom these variables have been assessed at study baseline, as well as their longitudinal coverage (i.e., assessment frequency and the number of participants assessed per visit) **(Fig. 4B)**. Additionally, links are provided that guide interested researchers directly to the data access applications of the respective datasets. The StudyPicker is particularly helpful for hypothesis-driven research or validation studies in which the variables that are elementary to conduct the planned analysis are often known in advance.

### 3.3 Detailed exploration of dataset content through interactive visualizations

Next to the semantic perspective, ADataViewer also allows for a detailed exploration of the integrated datasets based on descriptive statistics. Statistical distributions of numerical and categorical variables of interest can be visualized and compared across the available cohorts (https://adata.scai.fraunhofer.de/biomarkers). This functionality enables comparisons between individual diagnosis groups (i.e., cognitively unimpared (CU), mild cognitive impairment (MCI), AD) as well as the complete cohorts. Using these visualizations, researchers can investigate distributions and value representations encountered in the datasets and identify possible differences among them before starting their analysis.

A longitudinal view of the data can be generated in the ‘Longitudinal’ section. Dedicated visualizations display the follow-up per cohort on a variable-level **(Fig. 2)**.

**Figure 2:**
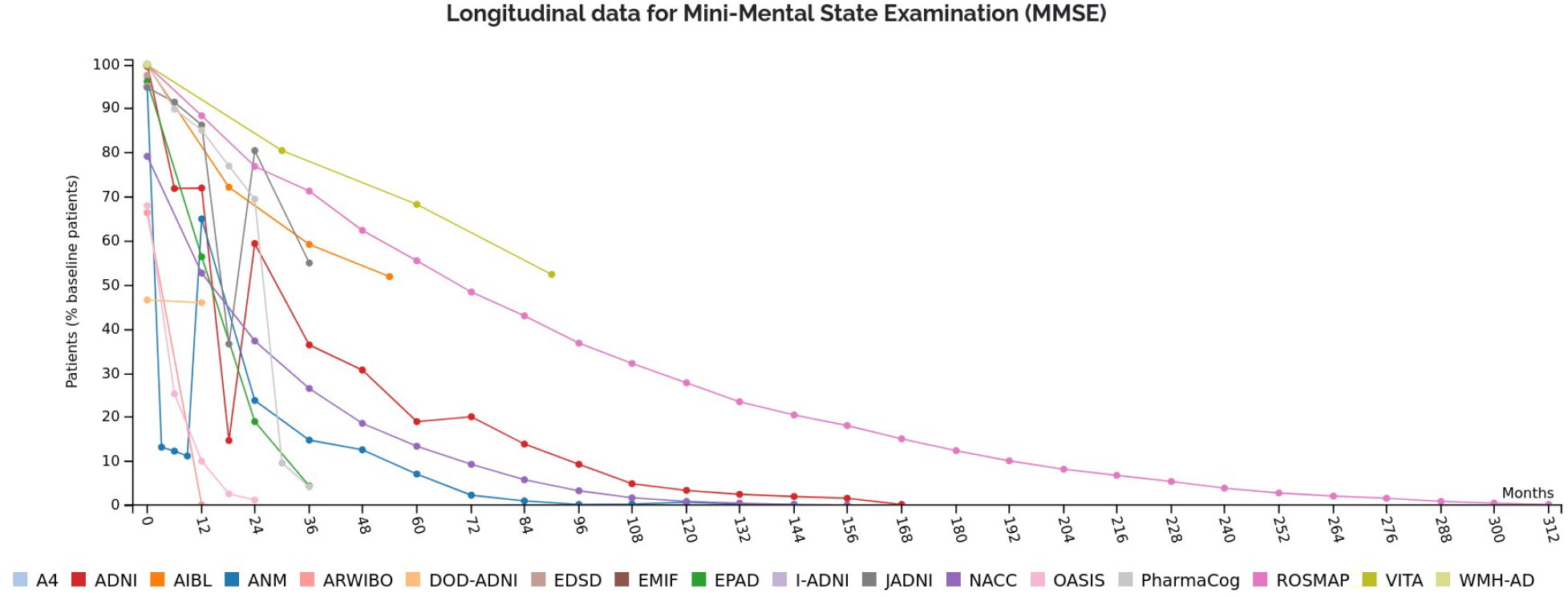
Exemplary longitudinal plot of the MMSE assessments generated using ADataViewer. Displayed are cohorts and their respective number of assessed participants for the selected variable.

### 3.4 Meta-analysis of cohort study content, assessed variables and common modalities

Besides the exploration and comparison of specific cohorts, ADataViewer helps to get a comprehensive overview on the state of the data landscape formed by the underlying cohorts. Here, the modality map (https://adata.scai.fraunhofer.de/modality) displays how commonly specific data modalities were included in cohort studies and, simultaneously, highlights areas that currently remain underexplored. Along the same line, **Fig. 3** shows an excerpt from an interactive visualization which depicts how many studies measured each individual variable. Furthermore, the plots displaying the ethnoracial diversity encountered in each individual cohort, and across cohorts grouped by geographic location, reveals over and underrepresentation of ethnoracial groups in data-driven AD research. All of this information can be vital when designing a novel cohort study aiming for either compatibility to other studies or at illuminating blind spots previously underrepresented in the AD data landscape.

**Figure 3:**
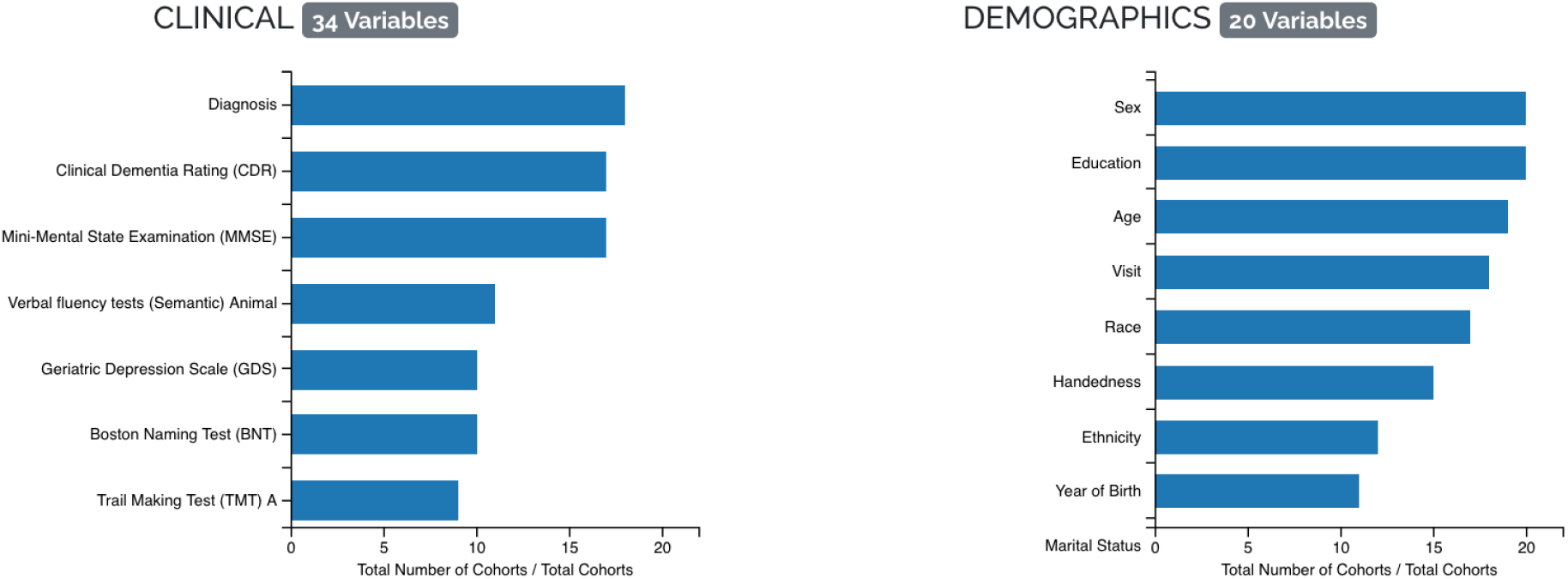
Excerpt of an interactive figure displaying the number of studies in which each specific variable was encountered (https://adata.scai.fraunhofer.de/biomarkers).

### 3.5 Exemplary application scenarios employing ADataViewer

#### Scenario 1

A researcher is searching for a discovery and validation cohort to model cognitive decline in the light of hippocampus atrophy, amyloid PET and depression. The variables of interest are: the Mini-Mental State Examination (MMSE), Clinical Dementia Rating Sum of Boxes (CDRSB), Hippocampus Volume, amyloid positron emission tomography (AV PET), Geriatric Depression Scale (GDS) and variables to correct for possible confounding (age, biological sex, education and APOE ε4 allele presence).

Given such a set of variables of interest, the StudyPicker of ADataViewer is the appropriate starting point to identify relevant cohorts. After submitting the variable query, we can directly observe that NACC, A4, ADNI, and DOD-ADNI contain all specified variables of interest **(Fig. 4A)**. However, after inspecting the follow-up plots, it is revealed that only NACC and ADNI hold sufficient longitudinal data to detect time-dependent relationships (here, 463 and 557 patients over 24 months study runtime, respectively) (**Fig. 4B and Fig. S1)**. Besides these two cohorts, EPAD, including 1,845 participants, could also provide a rich basis for the planned analysis if AV PET would be omitted **(Fig. 4A)**.

**Figure 4:**
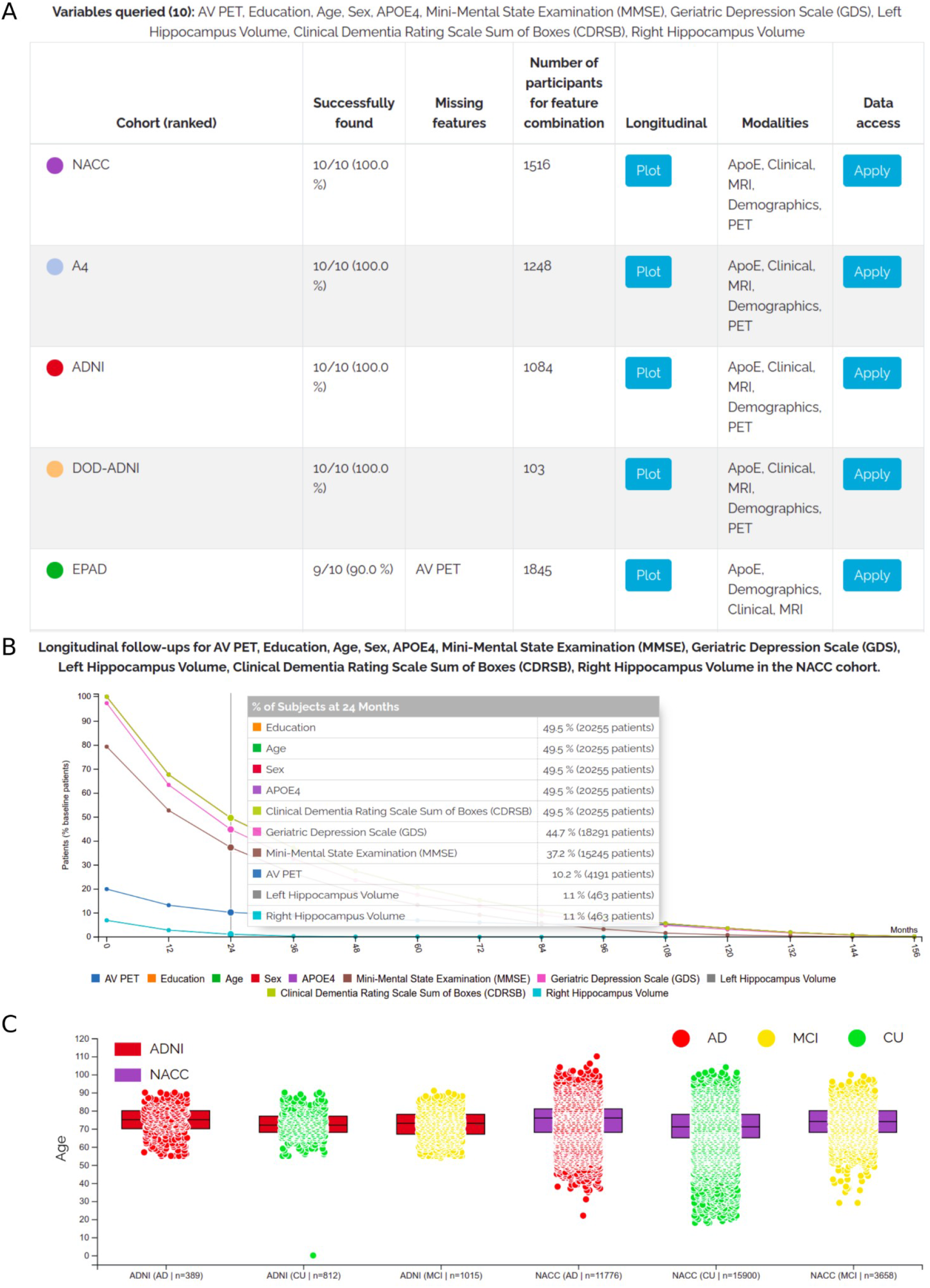
Case scenario of using ADataViewer to identify suitable cohort datasets with the aim to evaluate cognitive decline in the light of depression, AV PET and hippocampal atrophy. All graphs were created using the tools of ADataViewer. A, Excerpt of the ranking received by entering the variables of interest specified in application scenario 1 into the StudyPicker. B, Longitudinal coverage of the specified variables in the NACC cohort. See **Fig. S1** for the other cohorts’ plots. C, Comparison of the age distributions encountered across diagnostic groups of ADNI and NACC.

For a final evaluation on whether NACC and ADNI would suit the study needs, the ‘Biomarkers’ section can be used to compare cohort demographics and variable distributions. For example, comparing the age of participants in NACC and ADNI reveals a higher variance in the NACC data and presence of younger participants who would have been excluded from the ADNI study **(Fig. 4C)**. Furthermore, investigating the hippocampal volumes exposes a difference in value representation between the cohorts, as NACC values have been reported as normalized values **(Fig. S2)**. Consequently, it could be concluded that both datasets could be viable options for the discovery and replication process of a data-driven study, given that the representations of the hippocampal volume can be unified. Finally, the application process for data access can be initiated directly through the StudyPicker.

#### Scenario 2

A consortium is planning to conduct a longitudinal cohort study that aims at investigating AD in previously underrepresented ethnoracial groups. The assessed variables, however, should be compatible with other landmark AD cohorts to allow for a comparison of achieved results.

First, the ethnoracial diversity encountered across previous AD cohorts can be explored in the ‘Ethnicity’ section of ADataViewer. Their investigation demonstrates that 19 of the 20 cohorts enrolled predominantly caucasian/white participants. Keeping our proposed study goals in mind, it would therefore make sense to exclude caucasian/white participants from the recruitment of the envisioned study to focus on the currently underrepresented groups.

To achieve high compatibility with previous AD studies, the planned study should align its follow-up intervals and the assessed variables / data modalities to them. Here, the data modality map indicates that we should include demographics, clinical assessments, MRI, cerebrospinal fluid (CSF) biomarkers, at least APOE genotyping, administered medication, comorbidities, and the family history of participants to achieve a strong overlap in data modalities **(Fig. S3)**. More specifically, the most prominently assessed variables per modality can be explored in the ‘Biomarkers’ section **(Fig. 3)**. For example, we can observe that Clinical Dementia Rating (CDR), and MMSE are the most conducted cognitive assessments; demographics most commonly cover the biological sex, age, years of education, and ethnoracial group of participants; and phosphorylated tau, total tau and beta-amyloid were abundantly measured as CSF markers. By leveraging this information, we can make an informed decision on the variables we want to measure in the envisioned cohort study, such that an exploration of AD progression is feasible and that possible differences to cohorts of other ethnoracial compositions can be systematically evaluated. Additionally, the value ranges commonly encountered per variable can be explored using the biomarker boxplots **(Fig. 4C)**. Once the cohort study was conducted, we can use the provided variable mapping catalog to harmonize the new cohort dataset to all 20 datasets currently present in ADataViewer.

## 4. Discussion

ADataViewer aims at advancing patient data-driven AD research by increasing the findability and interoperability of cohort datasets and providing a deeper understanding of their content, both from a semantic and statistical perspective. The platform supports variable-level exploration of 20 AD cohort datasets and enables researchers to identify datasets suited for their envisioned studies before spending time on data access applications. In this context, we created, to the best of our knowledge, the most comprehensive variable mapping catalog in the AD domain that semantically harmonizes 1,196 unique variables across all investigated cohorts.

Following the FAIR data paradigm (findable, accessible, interoperable, reusable) [36], ADataViewer increases the findability of AD cohort datasets by displaying and suggesting possible data resources to researchers, enables better accessibility through direct links to the respective data access points, provides the variable mapping catalog to establish data interoperability, and facilitates the reuse of data for validation purposes. We believe that the presented platform can elevate data-driven AD research to be faster and more robust, because it becomes significantly easier to access the right datasets and validate results across multiple independent cohorts. In turn, this will help to better understand the heterogeneity across AD patients [37] and help to reveal possible cohort-specific findings [10].

Collecting patient-level data is a vastly expensive process. Therefore, studies are often limited with respect to their sample size, follow-up time, and variety of assessed data modalities. ADataViewer transparently provides researchers with information about what they can expect from specific datasets and whether it makes sense for them to spend a substantial amount of time on the acquisition of the individual data resource. Limiting the time spent on unfruitful dataset acquisitions will accelerate and benefit the actual analysis of the data. On this note, we would like to emphasize that ADataViewer is not meant to promote only the largest, most complete cohorts, but to show all available datasets that contain the information of interest for a conceived project. While larger cohorts often fare better as discovery cohorts, any cohort with equivalent information, regardless the size, could present a valuable resource for the subsequent validation of results and should therefore be considered.

### Limitations

One strength and simultaneous limitation of this work was its overarching premise that the data investigation was not based purely on descriptive metadata but on the dataset that was factually shared with us. Therefore, all results are based on the status of the distributed data and could vary from the content mentioned in official study reports or other versions of the same dataset. Ultimately, however, what drives the advancement of AD research is the factually shared, analyzable data and not what could potentially be available in theory.

The decision on how strict equivalence of variables is defined inevitably remains arbitrary to some degree. Here, we define two variables as semantically equivalent if the same information is presented in principle (i.e., the content of both variables can at least be broken down into the same information, see **Supplementary Material** for examples). Therefore, the acquisition method (e.g., type of MRI scanner) between two variables that were declared to be semantically equivalent may still differ and subsequent pre-processing of the raw data might be necessary to account for resulting statistical differences (e.g., elimination of batch effects). Sharing statistically harmonized data via ADataViewer is infeasible due to legal data sharing restrictions. However, the presented semantic mapping catalog presents a starting point to directly identify equivalent variables of interest and initiate the following pre-processing steps.

## 5. Conclusion

With ADataViewer, we aim to contribute to a robust, data-driven research culture that carefully reproduces and validates scientific results across multiple comparable datasets. As such, instead of pointing towards a single data resource, ADataViewer transparently displays the content of all integrated AD cohort datasets and the StudyPicker proposes all of these resources that match the researcher’s requirements. Our provided variable mappings build the basis for in-depth dataset comparisons and can act as a starting point to select and harmonize suited discovery and validation datasets.

## Data Availability

All reported results are available at https://adata.scai.fraunhofer.de. The datasets used in this publication are proprietary datasets owned by the respective data owners, access to the data can be requested using the information in https://adata.scai.fraunhofer.de/cohorts.

## Acknowledgements

We want to commend all data owners on their adherence to open science principles by sharing their data. We believe that their commitment is invaluable for any scientific research.

We thank the study participants and staff of the Rush Alzheimer’s Disease Center. ROSMAP was supported by NIA grants P30AG010161, R01AG015819, and R01AG017917.

The A4 Study is a secondary prevention trial in preclinical Alzheimer’s disease, aiming to slow cognitive decline associated with brain amyloid accumulation in clinically normal older individuals. The A4 Study is funded by a public-private-philanthropic partnership, including funding from the National Institutes of Health-National Institute on Aging, Eli Lilly and Company, Alzheimer’s Association, Accelerating Medicines Partnership, GHR Foundation, an anonymous foundation and additional private donors, with in-kind support from Avid and Cogstate. The companion observational Longitudinal Evaluation of Amyloid Risk and Neurodegeneration (LEARN) Study is funded by the Alzheimer’s Association and GHR Foundation. The A4 and LEARN Studies are led by Dr. Reisa Sperling at Brigham and Women’s Hospital, Harvard Medical School and Dr. Paul Aisen at the Alzheimer’s Therapeutic Research Institute (ATRI), University of Southern California. The A4 and LEARN Studies are coordinated by ATRI at the University of Southern California, and the data are made available through the Laboratory for Neuro Imaging at the University of Southern California. The participants screening for the A4 Study provided permission to share their de-identified data in order to advance the quest to find a successful treatment for Alzheimer’s disease. We would like to acknowledge the dedication of all the participants, the site personnel, and all of the partnership team members who continue to make the A4 and LEARN Studies possible. The complete A4 Study Team list is available on: a4study.org/a4-study-team.

Data collection and sharing of ABVIB was funded by the National Institutes on Aging (NIA) P01 AG12435.

Data collection and sharing of ARWIBO was supported by the Italian Ministry of Health, under the following grant agreements: Ricerca Corrente IRCCS Fatebenefratelli, Linea di Ricerca 2; Progetto Finalizzato Strategico 2000-2001 “Archivio normativo italiano di morfometria cerebrale con risonanza magnetica (età 40+)”; Progetto Finalizzato Strategico 2000-2001 “Decadimento cognitivo lieve non dementigeno: stadio preclinico di malattia di Alzheimer e demenza vascolare. Caratterizzazione clinica, strumentale, genetica e neurobiologica e sviluppo di criteri diagnostici utilizzabili nella realtà nazionale,”; Progetto Finalizzata 2002 “Sviluppo di indicatori di danno cerebrovascolare clinicamente significativo alla risonanza magnetica strutturale”; Progetto Fondazione CARIPLO 2005-2007 “Geni di suscettibilità per gli endofenotipi associati a malattie psichiatriche e dementigene”; “Fitness and Solidarietà”; and anonymous donors.

EPAD LCS is registered at www.clinicaltrials.gov Identifier: NCT02804789. Data used in preparation of this article were obtained from the EPAD LCS data set V.IMI, doi:10.34688/epadlcs_v.imi_20.10.30. The EPAD LCS was launched in 2015 as a public private partnership, led by Chief Investigator Professor Craig Ritchie MB BS. The primary research goal of the EPAD LCS is to provide a well-phenotyped probability-spectrum population for developing and continuously improving disease models for Alzheimer’s disease in individuals without dementia. This work used data and/or samples from the EPAD project which received support from the EU/EFPIA Innovative Medicines Initiative Joint Undertaking EPAD grant agreement n° 115736 and an Alzheimer’s Association Grant (SG21-818099-EPAD).

PharmaCog was funded through the European Community’s ‘Seventh Framework’ Programme (FP7/2007-2013) for an innovative scheme, the Innovative Medicines Initiative (IMI). IMI is a young and unique public-private partnership, founded in 2008 by the pharmaceutical industry (represented by the European Federation of Pharmaceutical Industries and Associations), EFPIA and the European Communities (represented by the European Commission).

J-A DNI was supported by the following grants: Translational Research Promotion Project from the New Energy and Industrial Technology Development Organization of Japan; Research on Dementia, Health Labor Sciences Research Grant; Life Science Database Integration Project of Japan Science and Technology Agency; Research Association of Biotechnology (contributed by Astellas Pharma Inc., Bristol-Myers Squibb, Daiichi-Sankyo, Eisai, Eli Lilly and Company, Merck-Banyu, Mitsubishi Tanabe Pharma, Pfizer Inc., Shionogi & Co., Ltd., Sumitomo Dainippon, and Takeda Pharmaceutical Company), Japan, and a grant from an anonymous Foundation.

Data collection and sharing for this project was funded by the Alzheimer’s Disease Neuroimaging Initiative (ADNI) (National Institutes of Health Grant U01 AG024904) and DOD ADNI (Department of Defense award number W81XWH-12-2-0012). ADNI is funded by the National Institute on Aging, the National Institute of Biomedical Imaging and Bioengineering, and through generous contributions from the following: AbbVie, Alzheimer’s Association; Alzheimer’s Drug Discovery Foundation; Araclon Biotech; BioClinica, Inc.; Biogen; Bristol-Myers Squibb Company; CereSpir, Inc.; Cogstate; Eisai Inc.; Elan Pharmaceuticals, Inc.; Eli Lilly and Company; EuroImmun; F. Hoffmann-La Roche Ltd and its affiliated company Genentech, Inc.; Fujirebio; GE Healthcare; IXICO Ltd.; Janssen Alzheimer Immunotherapy Research & Development, LLC.; Johnson & Johnson Pharmaceutical Research & Development LLC.; Lumosity; Lundbeck; Merck & Co., Inc.; Meso Scale Diagnostics, LLC.; NeuroRx Research; Neurotrack Technologies; Novartis Pharmaceuticals Corporation; Pfizer Inc.; Piramal Imaging; Servier; Takeda Pharmaceutical Company; and Transition Therapeutics. The Canadian Institutes of Health Research is providing funds to support ADNI clinical sites in Canada. Private sector contributions are facilitated by the Foundation for the National Institutes of Health (www.fnih.org). The grantee organization is the Northern California Institute for Research and Education, and the study is coordinated by the Alzheimer’s Therapeutic Research Institute at the University of Southern California. ADNI data are disseminated by the Laboratory for Neuro Imaging at the University of Southern California. This research was also supported by NIH grants P30 AG010129 and K01 AG030514.

The NACC database is funded by NIA/NIH Grant U01 AG016976. NACC data are contributed by the NIA-funded ADCs: P30 AG019610 (PI Eric Reiman, MD), P30 AG013846 (PI Neil Kowall, MD), P30 AG062428-01 (PI James Leverenz, MD) P50 AG008702 (PI Scott Small, MD), P50 AG025688 (PI Allan Levey, MD, PhD), P50 AG047266 (PI Todd Golde, MD, PhD), P30 AG010133 (PI Andrew Saykin, PsyD), P50 AG005146 (PI Marilyn Albert, PhD), P30 AG062421-01 (PI Bradley Hyman, MD, PhD), P30 AG062422-01 (PI Ronald Petersen, MD, PhD), P50 AG005138 (PI Mary Sano, PhD), P30 AG008051 (PI Thomas Wisniewski, MD), P30 AG013854 (PI Robert Vassar, PhD), P30 AG008017 (PI Jeffrey Kaye, MD), P30 AG010161 (PI David Bennett, MD), P50 AG047366 (PI Victor Henderson, MD, MS), P30 AG010129 (PI Charles DeCarli, MD), P50 AG016573 (PI Frank LaFerla, PhD), P30 AG062429-01(PI James Brewer, MD, PhD), P50 AG023501 (PI Bruce Miller, MD), P30 AG035982 (PI Russell Swerdlow, MD), P30 AG028383 (PI Linda Van Eldik, PhD), P30 AG053760 (PI Henry Paulson, MD, PhD), P30 AG010124 (PI John Trojanowski, MD, PhD), P50 AG005133 (PI Oscar Lopez, MD), P50 AG005142 (PI Helena Chui, MD), P30 AG012300 (PI Roger Rosenberg, MD), P30 AG049638 (PI Suzanne Craft, PhD), P50 AG005136 (PI Thomas Grabowski, MD), P30 AG062715-01 (PI Sanjay Asthana, MD, FRCP), P50 AG005681 (PI John Morris, MD), P50 AG047270 (PI Stephen Strittmatter, MD, PhD).

The results published here are in whole or in part based on data obtained from the AD Knowledge Portal (https://adknowledgeportal.org). These data are from the Vascular Contributors to Prodromal Alzheimer’s disease (VASCULAR) study. The study is part of the M2OVE-AD supported studies and is supported by NIH/NIA (PI: Ihab Hajjar, MD, MS grant numbers: AG051633 and AG057470. The investigators acknowledge the contribution of the study participants who donated their time and effort for this study.

## Author Contributions

CB conceived and supervised the project. YS and CB collected the datasets. YS prepared the data for ADataViewer. DDF implemented the platform. YS and CBA curated the variable mappings. CB drafted the manuscript. DDF, YS, and MHA revised the manuscript. MHA acquired the funding.

## References

1. Weiner, M. W., Veitch, D. P., Aisen, P. S., Beckett, L. A., Cairns, N. J., Cedarbaum, J., … & Alzheimer’s Disease Neuroimaging Initiative. (2015). Impact of the Alzheimer’s disease neuroimaging initiative, 2004 to 2014. Alzheimer’s & Dementia, 11(7), 865–884.

2. Shi, L., Westwood, S., Baird, A. L., Winchester, L., Dobricic, V., Kilpert, F., … & Nevado-Holgado, A. J. (2019). Discovery and validation of plasma proteomic biomarkers relating to brain amyloid burden by SOMAscan assay. Alzheimer’s & Dementia, 15(11), 1478–1488.

3. Koval, I., Bône, A., Louis, M., Lartigue, T., Bottani, S., Marcoux, A., … & Durrleman, S. (2021). AD Course Map charts Alzheimer’s disease progression. Scientific Reports, 11(1), 1–16.

4. Vogel, J. W., Young, A. L., Oxtoby, N. P., Smith, R., Ossenkoppele, R., Strandberg, O. T., … & Hansson, O. (2021). Four distinct trajectories of tau deposition identified in Alzheimer’s disease. Nature Medicine, 27(5), 871–881.

5. Fröhlich, H., Balling, R., Beerenwinkel, N., Kohlbacher, O., Kumar, S., Lengauer, T., … & Zupan, B. (2018). From hype to reality: data science enabling personalized medicine. BMC medicine, 16(1), 1–15.

6. Golriz Khatami, S., Robinson, C., Birkenbihl, C., Domingo-Fernández, D., Hoyt, C. T., & Hofmann-Apitius, M. (2020). Challenges of integrative disease modeling in Alzheimer’s disease. Frontiers in molecular biosciences, 6, 158.

7. Cunningham, J. A., Van Speybroeck, M., Kalra, D., & Verbeeck, R. (2016). Nine principles of semantic harmonization. In AMIA Annual Symposium Proceedings (Vol. 2016, p. 451). American Medical Informatics Association.

8. Neville, J., Kopko, S., Romero, K., Corrigan, B., Stafford, B., LeRoy, E., … & Stephenson, D. (2017). Accelerating drug development for Alzheimer’s disease through the use of data standards. Alzheimer’s & Dementia: Translational Research & Clinical Interventions, 3(2), 273–283.

9. Birkenbihl, C., Emon, M. A., Vrooman, H., Westwood, S., Lovestone, S., Hofmann-Apitius, M., & Fröhlich, H. (2020). Differences in cohort study data affect external validation of artificial intelligence models for predictive diagnostics of dementia-lessons for translation into clinical practice. EPMA Journal, 11(3), 367–376.

10. Birkenbihl, C., Salimi, Y., Fröhlich, H., Japanese Alzheimer’s Disease Neuroimaging Initiative, & Alzheimer’s Disease Neuroimaging Initiative. (2021). Unraveling the heterogeneity in Alzheimer’s disease progression across multiple cohorts and the implications for data-driven disease modeling. Alzheimer’s & Dementia.

11. Lovestone, S., & EMIF Consortium. (2020). The European medical information framework: a novel ecosystem for sharing healthcare data across Europe. Learning health systems, 4(2), e10214.

12. Gallacher, J., de Reydet de Vulpillieres, F., Amzal, B., Angehrn, Z., Bexelius, C., Bintener, C., … & ROADMAP Consortium. (2019). Challenges for optimizing real-world evidence in Alzheimer’s disease: the ROADMAP Project. Journal of Alzheimer’s Disease, 67(2), 495–501.

13. Oliveira, J. L., Trifan, A., & Silva, L. A. B. (2019). EMIF Catalogue: a collaborative platform for sharing and reusing biomedical data. International journal of medical informatics, 126, 35–45.

14. Janssen, O., Vos, S. J., García-Negredo, G., Tochel, C., Gustavsson, A., Smith, M., … & Diaz, C. (2020). Real-world evidence in Alzheimer’s disease: the ROADMAP Data Cube. Alzheimer’s & Dementia, 16(3), 461–471.

15. Birkenbihl, C., Salimi, Y., Domingo-Fernándéz, D., Lovestone, S., AddNeuroMed consortium, Fröhlich, H., … & Alzheimer’s Disease Neuroimaging Initiative. (2020). Evaluating the Alzheimer’s disease data landscape. Alzheimer’s & Dementia: Translational Research & Clinical Interventions, 6(1), e12102.

16. Sperling, R. A., Rentz, D. M., Johnson, K. A., Karlawish, J., Donohue, M., Salmon, D. P., & Aisen, P. (2014). The A4 study: stopping AD before symptoms begin?. Science translational medicine, 6(228), 228fs13–228fs13.

17. Rodriguez, F. S., Zheng, L., Chui, H. C., & Aging Brain: Vasculature, Ischemia, and Behavior Study (2019). Psychometric Characteristics of Cognitive Reserve: How High Education Might Improve Certain Cognitive Abilities in Aging. Dementia and geriatric cognitive disorders, 47(4-6), 335–344. https://doi.org/10.1159/000501150.

18. Mueller, S. G., Weiner, M. W., Thal, L. J., Petersen, R. C., Jack, C. R., Jagust, W., … & Beckett, L. (2005). Ways toward an early diagnosis in Alzheimer’s disease: the Alzheimer’s Disease Neuroimaging Initiative (ADNI). Alzheimer’s & Dementia, 1(1), 55–66.

19. Ellis, K. A., Bush, A. I., Darby, D., De Fazio, D., Foster, J., Hudson, P., … & AIBL Research Group. (2009). The Australian Imaging, Biomarkers and Lifestyle (AIBL) study of aging: methodology and baseline characteristics of 1112 individuals recruited for a longitudinal study of Alzheimer’s disease. International psychogeriatrics, 21(4), 672–687.

20. Birkenbihl, C., Westwood, S., Shi, L., Nevado-Holgado, A., Westman, E., Lovestone, S., … & AddNeuroMed Consortium. (2020). ANMerge: a comprehensive and accessible Alzheimer’s disease patient-level dataset. Journal of Alzheimer’s Disease, (Preprint), 1–9.

21. Frisoni, G. B., Prestia, A., Zanetti, O., Galluzzi, S., Romano, M., Cotelli, M., … & Geroldi, C. (2009). Markers of Alzheimer’s disease in a population attending a memory clinic. Alzheimer’s & Dementia, 5(4), 307–317.

22. Weiner, M. W., Veitch, D. P., Hayes, J., Neylan, T., Grafman, J., Aisen, P. S., … & Department of Defense Alzheimer’s Disease Neuroimaging Initiative. (2014). Effects of traumatic brain injury and posttraumatic stress disorder on Alzheimer’s disease in veterans, using the Alzheimer’s Disease Neuroimaging Initiative. Alzheimer’s & dementia, 10, S226–S235.

23. Brueggen, K., Grothe, M. J., Dyrba, M., Fellgiebel, A., Fischer, F., Filippi, M., … & Teipel, S. (2017). The European DTI Study on Dementia—a multicenter DTI and MRI study on Alzheimer’s disease and mild cognitive impairment. NeuroImage, 144, 305–308.

24. Bos, I., Vos, S., Vandenberghe, R., Scheltens, P., Engelborghs, S., Frisoni, G., … & Visser, P. J. (2018). The EMIF-AD Multimodal Biomarker Discovery study: design, methods and cohort characteristics. Alzheimer’s research & therapy, 10(1), 1–9.

25. Solomon, A., Kivipelto, M., Molinuevo, J. L., Tom, B., & Ritchie, C. W. (2018). European prevention of Alzheimer’s dementia longitudinal cohort study (EPAD LCS): study protocol. BMJ open, 8(12), e021017.

26. Cavedo, E., Redolfi, A., Angeloni, F., Babiloni, C., Lizio, R., Chiapparini, L., Bruzzone, M. G., Aquino, D., Sabatini, U., Alesiani, M., Cherubini, A., Salvatore, E., Soricelli, A., Vernieri, F., Scrascia, F., Sinforiani, E., Chiarati, P., Bastianello, S., Montella, P., Corbo, D., … Frisoni, G. B. (2014). The Italian Alzheimer’s Disease Neuroimaging Initiative (I-ADNI): validation of structural MR imaging. Journal of Alzheimer’s disease : JAD, 40(4), 941–952. https://doi.org/10.3233/JAD-132666.

27. Iwatsubo, T. (2010). Japanese Alzheimer’s Disease Neuroimaging Initiative: present status and future. Alzheimer’s & Dementia, 6(3), 297–299.

28. Besser, L., Kukull, W., Knopman, D. S., Chui, H., Galasko, D., Weintraub, S., … & Morris, J. C. (2018). Version 3 of the National Alzheimer’s Coordinating Center’s Uniform Data Set. Alzheimer disease and associated disorders.

29. Marcus, D. S., Fotenos, A. F., Csernansky, J. G., Morris, J. C., & Buckner, R. L. (2010). Open access series of imaging studies: longitudinal MRI data in nondemented and demented older adults. Journal of cognitive neuroscience, 22(12), 2677–2684.

30. Breitner, J. C. S., Poirier, J., Etienne, P. E., & Leoutsakos, J. M. (2016). Rationale and Structure for a New Center for Studies on Prevention of Alzheimer’s Disease (StoP-AD). The journal of prevention of Alzheimer’s disease, 3(4), 236–242.

31. Galluzzi, S., Marizzoni, M., Babiloni, C., Albani, D., Antelmi, L., Bagnoli, C., Bartres-Faz, D., Cordone, S., Didic, M., Farotti, L., Fiedler, U., Forloni, G., Girtler, N., Hensch, T., Jovicich, J., Leeuwis, A., Marra, C., Molinuevo, J. L., Nobili, F., Pariente, J., … PharmaCog Consortium (2016). Clinical and biomarker profiling of prodromal Alzheimer’s disease in workpackage 5 of the Innovative Medicines Initiative PharmaCog project: a ‘European ADNI study’. Journal of internal medicine, 279(6), 576–591. https://doi.org/10.1111/joim.12482

32. A Bennett, D. A Schneider, J., Arvanitakis, Z., & S Wilson, R. (2012). Overview and findings from the religious orders study. Current Alzheimer Research, 9(6), 628–645.

33. Emory University School of Medicine (2021, July). VASCULAR (VAScular ContribUtors to prodromaL AlzheimeR’s disease). https://med.emory.edu/departments/medicine/divisions/geriatrics-gerontology/research/labs/bsharp/studies.html

34. Fischer, P., Jungwirth, S., Krampla, W., Weissgram, S., Kirchmeyr, W., Schreiber, W., … & Tragl, K. H. (2002). Vienna Transdanube Aging “VITA”: study design, recruitment strategies and level of participation. In Ageing and Dementia Current and Future Concepts (pp. 105–116). Springer, Vienna.

35. Damulina, A., Pirpamer, L., Seiler, S., Benke, T., Dal-Bianco, P., Ransmayr, G., … & Schmidt, R. (2019). White matter hyperintensities in Alzheimer’s disease: a lesion probability mapping study. Journal of Alzheimer’s Disease, 68(2), 789–796.

36. Wilkinson, M. D., Dumontier, M., Aalbersberg, I. J., Appleton, G., Axton, M., Baak, A., … & Mons, B. (2016). The FAIR Guiding Principles for scientific data management and stewardship. Scientific data, 3(1), 1–9.

37. Verdi, S., Marquand, A. F., Schott, J. M., & Cole, J. H. (2021). Beyond the average patient: how neuroimaging models can address heterogeneity in dementia. Brain.

