## Supplementary material for "ADataViewer: Exploring Semantically Harmonized Alzheimer’s Disease Cohort Datasets": Figure S1, Figure S2, Figure S3

### Variable namespace harmonization

While some variables could be mapped unambiguously between the datasets (e.g. the MMSE or other standardized clinical assessments), others led to a trade-off between semantic accuracy and a complete mapping. For example, given studies measured the smoking behaviour of participants, we could observe that the

* Alzheimer's Disease Neuroimaging Initiative: Data used in the preparation of this article were obtained from the Alzheimer's Disease Neuroimaging Initiative (ADNI) database (adni.loni.usc.edu). As such, the investigators within the ADNI contributed to the design and implementation of ADNI and/or provided data but did not participate in analysis or writing of this report. A complete listing of ADNI investigators can be found at http://adni.loni. usc.edu/wp-content/uploads/how_to_apply/ ADNI_Acknowledgement_List.pdf

Ж Japanese Alzheimer's Disease Neuroimaging Initiative: Data used in preparation of this article were obtained from the Japanese Alzheimer's Disease Neuroimaging Initiative (J-ADNI) database deposited in the National Bioscience Database Center Human Database, Japan (Research ID: hum0043.v1, 2016). As such, the investigators within J-ADNI contributed to the design and implementation of J-ADNI and/or provided data but did not participate in analysis or writing of this report. A complete listing of J-ADNI investigators can be found at: https://humandbs.biosciencedbc.jp/ en/hum0043-j-adni-authors.

Ψ Data used in preparation of this article were obtained from the Aging Brain: Vasculature, Ischemia, and Behavior Study (ABVIB). As such, the key investigators within the ABVIB contributed to the design and implementation of ABVIB and/or provided data but did not participate in analysis or writing of this report: Helena C. Chui M.D. (Principal Investigator), Charles C. DeCarli, M.D., William G. Ellis, M.D., William J. Jagust, M.D., Joel H. Kramer, Ph.D., Meng Law, M.D., Dan Mungas Ph,D. Bruce R. Reed, Ph.D., Nerses Sanossian, M.D., Michael W. Weiner, M.D. Wendy J. Mack, Ph.D., Harry V. Vinters, M.D., Chris Zarow, Ph.D., Ling Zheng, Ph.D.

‡ Data used in preparation of this article were obtained from the Alzheimer's Disease Repository Without Borders (ARWiBo) database (www.arwibo.it). As such, the researchers within the ARWiBo contributed to the design and implementation of ARWiBo and/or provided data but did not participate in analysis or writing of this report. A complete listing of ARWiBo researchers can be found at: www.arwibo.it/acknowledgement.it

† Data used in preparation of this article were obtained from the Longitudinal Cohort Study (LCS), delivered by the European Prevention of Alzheimer's Disease (EPAD) Consortium. As such investigators within the EPAD LCS and EPAD Consortium contributed to the design and implementation of EPAD and/or provided data but did not participate in analysis or writing of this report. A complete list of EPAD Investigators can be found at: http://ep-ad.org/wp-content/uploads/2020/12/202010_List-of-epadistas.pdf

corresponding information was reported in varying detail: study A would report a binary variable “smoking yes/no”, study B would provide ordinal information “participant smokes none/1-3 cigarretes/4-10 cigarretes/>10 cigarettes per day”, while study C might have posed the question whether participants “smoked at least one cigarette over the last week”. While these variables measured three different aspects of smoking in the strict sense, we still mapped them together as their information could be made equivalent through appropriate pre-processing (here, the largest common denominator between those variables would be a binary indication whether the participant smokes at least occasionally).

### Linking mapped variables to referential ontologies

During the mapping process, we also linked variables to referential ontologies with the goal of cross-reference these variables with existing controlled vocabularies. Thus, each of the final variables were queried against definitions present in any of the ontologies indexed by the Ontology Lookup Service (OLS) (https://www.ebi.ac.uk/ols/index). In case of multiple matches, we preferably linked the variables to referential ontologies as they are constantly updated and contain cross-references to similar vocabularies. In total, we linked the variables in ADataViewer to 7 distinct ontologies: Uber-anatomy Ontology (UBERON), Foundational Model of Anatomy Ontology (FMA), Ontology for MIRNA Target (OMIT), National Cancer Institute Thesaurus (NCIT), Experimental Factor Ontology (EFO), Human Phenotype (HP), and Mondo Disease Ontology (MONDO).

A **Longitudinal follow-ups for AV PET, Education, Age, Sex, APOE4, Mini-Mental State Examination (MMSE), Geriatric Depression Scale (GDS), Left Hippocampus Volume, Clinical Dementia Rating Scale Sum of Boxes (CDRSB), Right Hippocampus Volume in the NACC cohort.**

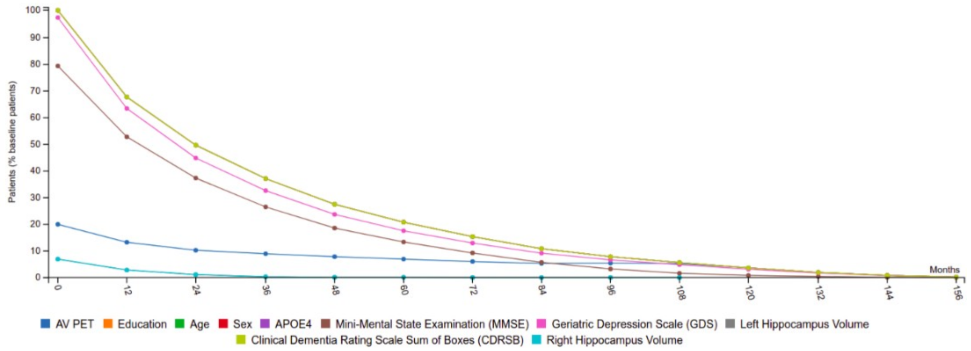


B **Longitudinal follow-ups for AV PET, Education, Age, Sex, APOE4, Mini-Mental State Examination (MMSE), Geriatric Depression Scale (GDS), Left Hippocampus Volume, Clinical Dementia Rating Scale Sum of Boxes (CDRSB), Right Hippocampus Volume in the A4 cohort.**

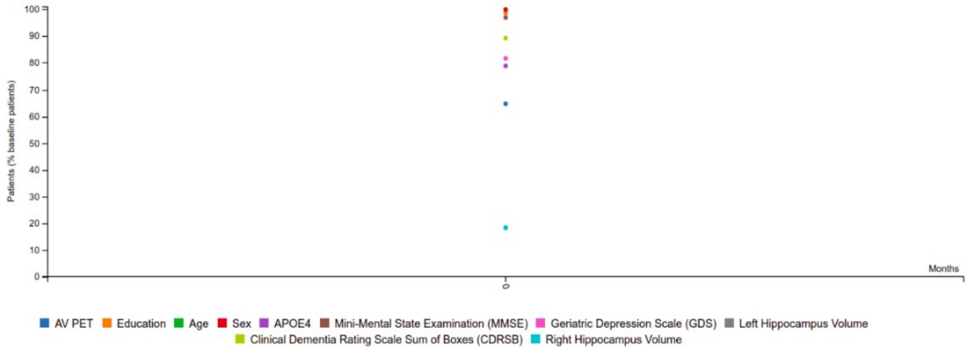


C **Longitudinal follow-ups for AV PET, Education, Age, Sex, APOE4, Mini-Mental State Examination (MMSE), Geriatric Depression Scale (GDS), Left Hippocampus Volume, Clinical Dementia Rating Scale Sum of Boxes (CDRSB), Right Hippocampus Volume in the ADNI cohort.**

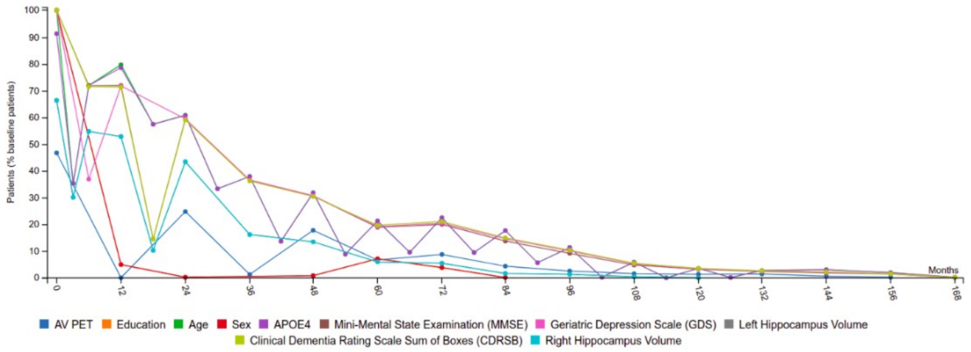


D **Longitudinal follow-ups for AV PET, Education, Age, Sex, APOE4, Mini-Mental State Examination (MMSE), Geriatric Depression Scale (GDS), Left Hippocampus Volume, Clinical Dementia Rating Scale Sum of Boxes (CDRSB), Right Hippocampus Volume in the DOD-ADNI cohort.**

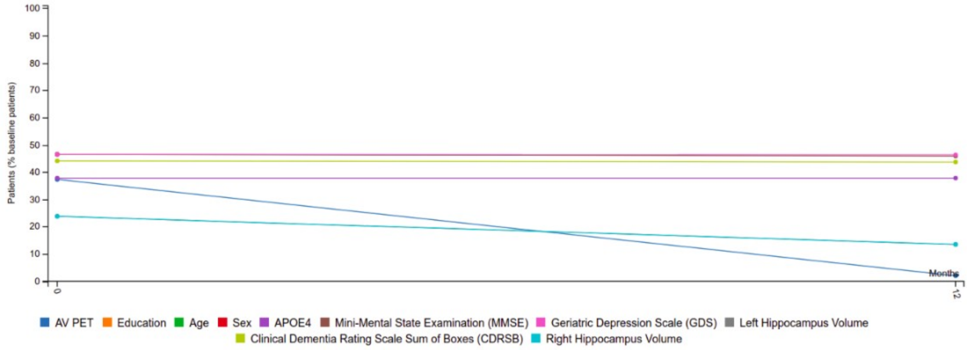

**Figure S1:** Longitudinal follow-up plots the specified variables corresponding to the StupyPicker application scenario for A, the NACC cohort. B, the A4 cohort. C, the ADNI cohort. D, the DOD-ADNI cohort.

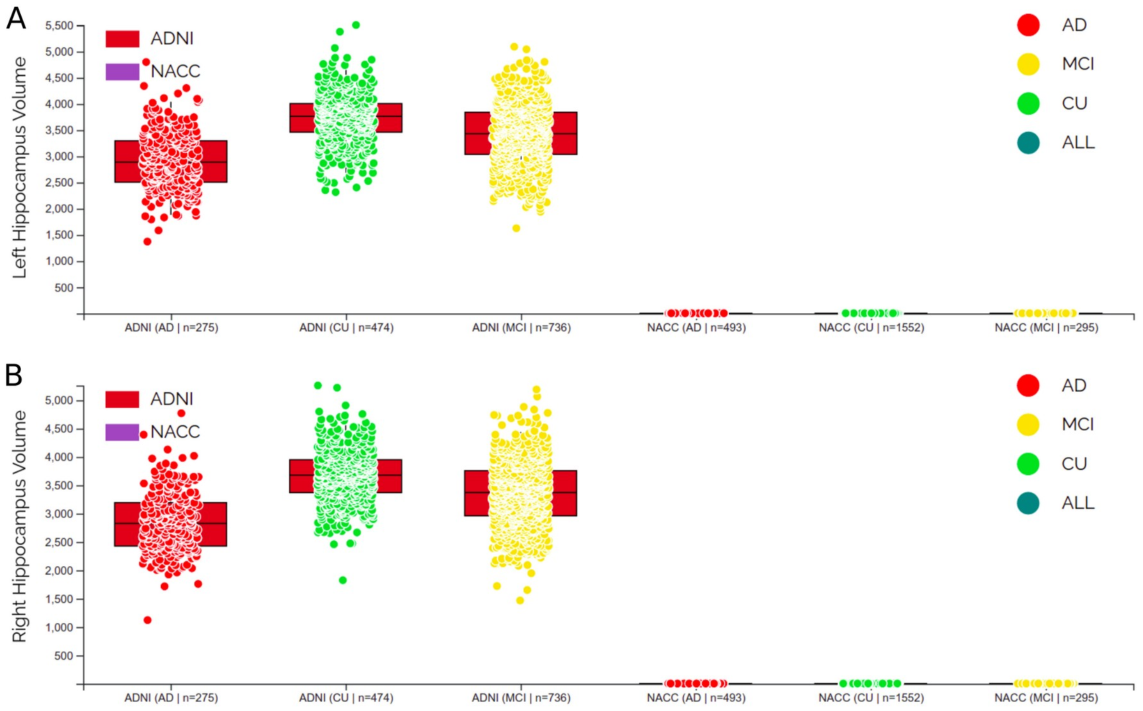


**Figure S2:** Distribution of hippocampus volume displayed with boxplots using the "Biomarkers" tool of the ADataViewer. A, Left hippocampus volume across diagnostic groups of ADNI and NACC. B, Right hippocampus volume across diagnostic groups of ADNI and NACC.

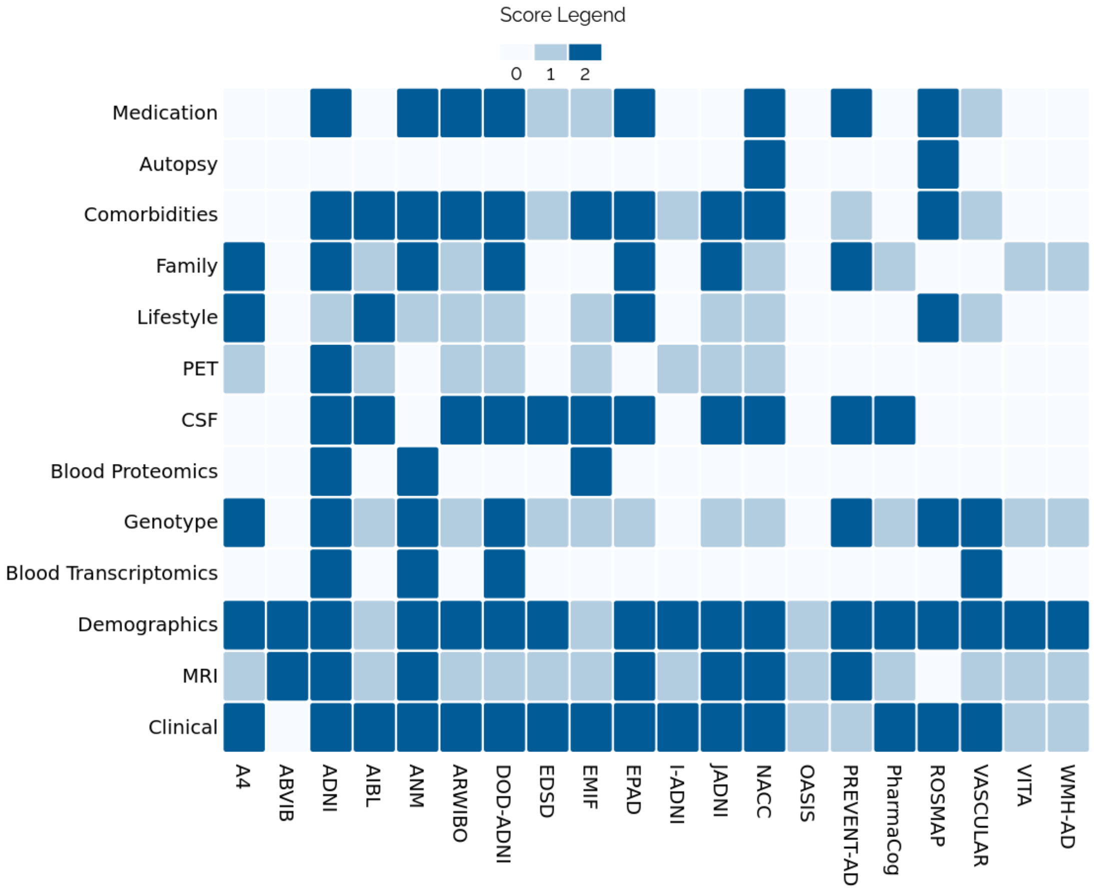


**Figure S3:** The modality map, describing which data modalities have been assessed per cohort. A detailed explanation of the scores is provided under https://adata.scai.fraunhofer.de/modality#criteria. In brief, a score of 0 means the modality was not measured, 1 means was partially measured and/or shared, 2 means it was available.
